# The relationship between neighborhood poverty and COVID-19 mortality within racial/ethnic groups (Cook County, Illinois)

**DOI:** 10.1101/2020.10.04.20206318

**Authors:** Justin M. Feldman, Mary T. Bassett

## Abstract

**Background:** Prior research has identified higher rates of COVID-19 mortality among people of color (relative to non-Hispanic whites) and populations in high-poverty neighborhoods (relative to wealthier neighborhoods). It is unclear, however, whether non-Hispanic whites in high-poverty neighborhoods experience elevated mortality, or whether people of color living in wealthy areas are relatively protected. Exploring socioeconomic position in combination with race/ethnicity can lead to a more detailed understanding of the specific processes that result in COVID-19 inequities.

**Methods and Findings:** We used census and individual-level mortality data for the non-Hispanic white, non-Hispanic Black, and Hispanic/Latinx populations of Cook County, Illinois, USA. We excluded deaths related to nursing homes and other institutions. We calculated age and gender-adjusted mortality rates by race/ethnicity, census tract poverty quartile, and age group (0-64 and ≥65 years).

Within all racial/ethnic groups, COVID-19 mortality rates were greatest in the highest-poverty quartile and lowest in the lowest-poverty quartile. The mortality rate for younger non-Hispanic whites in the highest-poverty quartile was 13.5 times that of younger non-Hispanic whites in the lowest-poverty quartile (95% CI: 8.5, 21.4). For young people in the highest-poverty quartile, the non-Hispanic white and Black mortality rates were similar. Among younger people in the lowest-poverty quartile, non-Hispanic Black and Hispanic/Latinx people had mortality rates nearly three times that of non-Hispanic whites. For the older population, the mortality rate among non-Hispanic whites in the highest-poverty quartile was less than that of lowest-poverty non-Hispanic Black and Hispanic/Latinx populations.

**Conclusions:** Our findings suggest racial/ethnic inequalities in COVID-19 mortality are partly, but not entirely, attributable to the higher average socioeconomic position of non-Hispanic whites relative to the non-Hispanic Black and Hispanic/Latinx populations. Future research on health equity in COVID-19 outcomes should collect and analyze individual-level data on the potential mechanisms driving population distributions of exposure, severe illness, and death.

---

Racial/ethnic inequities in US COVID-19 mortality are now widely documented: death rates are substantially higher among non-Hispanic Black (NHB), Hispanic/Latinx, and American Indian populations compared to non-Hispanic whites (NHWs).^1^ There is also evidence of socioeconomic gradients in COVID-19 mortality, meaning residents of economically deprived neighborhoods experience the highest mortality and those in the wealthiest neighborhoods experience the lowest.^2^ It is unclear, however, whether NHWs in high-poverty neighborhoods experience elevated mortality, or whether people of color living in wealthy areas are relatively protected. Discussions of racial/ethnic groups in relation to COVID-19 often lack context about the modifiable social and economic processes, rooted in structural racism, that lead to inequality.^3^ Exploring the role of socioeconomic position in COVID-19 mortality in combination with race/ethnicity can help lead to a more detailed understanding of the specific processes that result in health inequities.

There are a number of key limitations to prior research on socioeconomic gradients for COVID-19 mortality, particularly because individual-level mortality data from 2020 are still largely unavailable to researchers. Without such data, it is difficult to determine whether socioeconomic gradients exist within racial/ethnic groups and whether these gradients are similar across groups. Additionally, prior studies of socioeconomic gradients in COVID-19 outcomes have rarely distinguished between populations residing in institutions versus households. This is a concern because a high proportion of COVID-19 deaths (exceeding 50% in many states) occur among persons in nursing homes.^4^ While neighborhood-based measures such as poverty rates can serve as meaningful proxies for socioeconomic position for residents of households^5^, it is unclear whether the same is true for populations in intuitions (e.g. if nursing home’s location in a high-poverty area, it may not indicate that its residents tend to be economically deprived).

For COVID-19 mortality, one prior study found that the full (institutional and household) population exhibited a weaker social gradient than the household population alone.^6^ Analyses that do not differentiate between the two populations are likely capturing artifacts related to the geography of nursing home siting rather than the population distribution of COVID-19 by socioeconomic position.

This cross-sectional study characterizes race/ethnicity-specific socioeconomic differences in COVID-19 mortality for using a unique individual-level, open-access dataset on mortality in Cook County, Illinois (population = 5.2 million, about half of which resides in Chicago). It excludes deaths attributed to nursing homes and other institutions, restricting to deaths among the population in households. Informed by prior research on the population distributions of chronic conditions and mortality^7–10^, we hypothesized that COVID-19 mortality rates would exhibit socioeconomic gradients within each racial/ethnic group.

## Methods

We obtained data on deaths due to COVID-19 that were reported in the Cook County, Illinois Medical Examiner Case Archive as of September 14, 2020.^11^ The case archive includes cause of death fields along with demographic characteristics and geography, including an already geocoded “incident location” which, for COVID-19, corresponds to the place where the illness was first noted. We geocoded incident locations for which there was a non-missing street address but missing latitude and longitude. We identified deaths for which COVID-19 was indicated in one or more cause of death fields and had a race/ethnicity of NHW, NHB, or Hispanic/Latinx. There were too few deaths occurring among other racial/ethnic groups to be meaningfully analyzed.

The case archive did not have a field to indicate deaths linked to institutions. We identified these deaths for exclusion through (1) nearest-neighbor matching incident location centroids to skilled nursing facility centroids (using a distance cutoff of <250 meters) based on a statewide facility licensing database in Illinois, which contains data on licensed nursing homes and rehabilitation centers^12^; (2) flagging incident locations that included “nursing home”, “rehab”, or similar in the street address field, or that shared coordinates with such a location; and (3) investigating, via web searches, any incident location appearing three or more times and flagging it if it was a nursing home, rehabilitation center, assisted living home, or hospital (according to state data, 97% of nursing home-related COVID-19 deaths in Cook County occurred in nursing homes with at least 3 total deaths^13^).

We used the geographic coordinates for each non-institutional incident location to determine its census tract, a geographic unit akin to neighborhoods. The 2018 American Community Survey provided census tract poverty rates (% of the population living below the federal poverty level) and population denominators for rate calculations. Poverty quartiles ranged from >21.4% for tracts with the highest poverty rates to <6.6% for those with the lowest. We aggregated the data such that each row contained the sum of all deaths and the sum of the residential population for a given poverty quartile, racial/ethnic group, age category, and gender. We calculated adjusted mortality rates and 95% confidence intervals (95% CIs) for each racial/ethnic group and poverty quartile using direct standardization in Stata version 16 (Stata Corp., College Station, Texas), adjusting subgroups to the age and gender distribution of the full study population. We calculated mortality rate ratios and 95% CIs using standard methods,^14^ with analyses for the entire study population and also stratified by age group for the younger (0 to 64 years) and older (≥65 years) populations.

## Results

As of September 29, 2020, the Cook County, Illinois Medical Examiner reported 5,216 total deaths due to COVID-19. The first death occurred on March 16, 2020 and the most recent occurred on September 27, 2020. We excluded 2,880 deaths (414 were not geocoded with precision, 184 had incident locations outside of Cook County, 2,113 were associated with nursing homes or other institutions, 146 were an excluded or missing race/ethnicity, and 3 were missing age or gender), yielding 2,336 included deaths (Table 1a). Of included deaths, the mean age was 68 years (standard deviation = 15 years) and 62% were men.

**Table 1a.** Mortality due to COVID-19 in Cook County, Illinois by race/ethnicity and census tract poverty rate (Non-Hispanic Black, Non-Hispanic white, and Hispanic/Latinx persons). All ages, population in households.

| Population group | Census tract poverty quartile | COVID-19 deaths (n) | Population size (N) | % of population group residing in quartile | Adjusted mortality rate |  |  | Mortality rate ratio |  |  |
| --- | --- | --- | --- | --- | --- | --- | --- | --- | --- | --- |
|  |  |  |  |  | Rate per 100,000 | 95% CI (lower bound) | 95% CI (upper bound) | Rate ratio (vs. quartile 4) | 95% CI (lower bound) | 95% CI (upper bound) |
| All three racial/ethnic groups | Total | 2,336 | 4,762,460 | 100.0% | 49.0 | 47.1 | 51.0 |  |  |  |
|  | 1 (high poverty) | 918 | 1,186,847 | 24.9% | 91.5 | 85.6 | 97.4 | 4.5 | 3.9 | 5.1 |
|  | 2 | 702 | 1,190,873 | 25.0% | 63.7 | 59.0 | 68.4 | 3.1 | 2.7 | 3.6 |
|  | 3 | 424 | 1,190,708 | 25.0% | 34.0 | 30.8 | 37.2 | 1.7 | 1.4 | 1.9 |
|  | 4 (low poverty) | 292 | 1,194,032 | 25.1% | 20.4 | 18.0 | 22.7 | 1.0 | – | – |
| White, non-Hispanic | Total |  | 2,217,734 | 100.0% | 22.8 | 21.0 | 24.5 |  |  |  |
|  | 1 (high poverty) | 110 | 171,567 | 7.7% | 58.3 | 47.4 | 69.3 | 4.1 | 3.3 | 5.2 |
|  | 2 | 188 | 417,220 | 18.8% | 35.3 | 30.2 | 40.3 | 2.5 | 2.0 | 3.1 |
|  | 3 | 180 | 698,118 | 31.5% | 19.5 | 16.6 | 22.4 | 1.4 | 1.1 | 1.7 |
|  | 4 (low poverty) | 185 | 930,829 | 42.0% | 14.1 | 12.1 | 16.2 | 1.0 | – | – |
| Black, non-Hispanic | Total |  | 1,230,494 | 100.0% | 77.1 | 72.0 | 82.2 |  |  |  |
|  | 1 (high poverty) | 526 | 670,615 | 54.5% | 87.6 | 80.0 | 95.1 | 1.6 | 1.2 | 2.2 |
|  | 2 | 216 | 299,308 | 24.3% | 71.8 | 62.2 | 81.5 | 1.3 | 0.9 | 1.8 |
|  | 3 | 105 | 181,064 | 14.7% | 59.9 | 48.4 | 71.5 | 1.1 | 0.8 | 1.6 |
|  | 4 (low poverty) | 43 | 79,507 | 6.5% | 55.1 | 38.3 | 71.8 | 1.0 | – | – |
| Hispanic/Latinx | Total |  | 1,314,232 | 100.0% | 102.4 | 94.8 | 110.1 |  |  |  |
|  | 1 (high poverty) | 282 | 344,665 | 26.2% | 135.1 | 118.1 | 152.1 | 2.1 | 1.5 | 2.8 |
|  | 2 | 298 | 474,345 | 36.1% | 109.4 | 96.2 | 122.7 | 1.7 | 1.2 | 2.2 |
|  | 3 | 139 | 311,526 | 23.7% | 78.0 | 64.3 | 91.7 | 1.2 | 0.9 | 1.6 |
|  | 4 (low poverty) | 64 | 183,696 | 14.0% | 65.5 | 48.1 | 82.8 | 1.0 | – | – |

Lower bounds for census tract poverty rates by quartile: (1) 21.4%; (2) 12.3%; (3) 6.6%; (4) 0.0%.

**Table 1b.** Mortality due to COVID-19 in Cook County, Illinois by race/ethnicity and census tract poverty rate (Non-Hispanic Black, Non-Hispanic white, and Hispanic/Latinx persons). Ages 0 to 64, population in households.

| Population group | Census tract poverty quartile | COVID-19 deaths (n) | Population size (N) | % of population group residing in quartile | Adjusted mortality rate |  |  | Mortality rate ratio |  |  |
| --- | --- | --- | --- | --- | --- | --- | --- | --- | --- | --- |
|  |  |  |  |  | Rate per 100,000 | 95% CI (lower bound) | 95% CI (upper bound) | Rate ratio (vs. quartile 4) | 95% CI (lower bound) | 95% CI (upper bound) |
| All three racial/ethnic groups | Total | 883 | 4,091,496 | 100.0% | 21.6 | 20.2 | 23.0 |  |  |  |
|  | 1 (high poverty) | 394 | 1,048,231 | 25.6% | 42.5 | 38.3 | 46.8 | 6.6 | 5.1 | 8.5 |
|  | 2 | 273 | 1,037,906 | 25.4% | 27.5 | 24.3 | 30.8 | 4.3 | 3.3 | 5.5 |
|  | 3 | 143 | 1,013,042 | 24.8% | 13.8 | 11.5 | 16.0 | 2.1 | 1.6 | 2.8 |
|  | 4 (low poverty) | 73 | 992,317 | 24.3% | 6.5 | 5.0 | 8.0 | 1.0 | – | – |
| White, non-Hispanic | Total | 173 | 1,799,663 | 100.0% | 8.1 | 6.9 | 9.3 |  |  |  |
|  | 1 (high poverty) | 58 | 143,713 | 8.0% | 37.8 | 28.0 | 47.6 | 13.5 | 8.5 | 21.4 |
|  | 2 | 51 | 339,590 | 18.9% | 12.9 | 9.3 | 16.5 | 4.6 | 2.9 | 7.4 |
|  | 3 | 37 | 565,898 | 31.4% | 5.6 | 3.8 | 7.3 | 2.0 | 1.2 | 3.3 |
|  | 4 (low poverty) | 27 | 750,462 | 41.7% | 2.8 | 1.7 | 3.9 | 1.0 | – | – |
| Black, non-Hispanic | Total | 299 | 1,062,219 | 100.0% | 28.9 | 25.6 | 32.1 |  |  |  |
|  | 1 (high poverty) | 177 | 582,600 | 54.8% | 33.1 | 28.2 | 38.1 | 1.4 | 0.9 | 2.3 |
|  | 2 | 75 | 254,762 | 24.0% | 28.2 | 21.8 | 34.7 | 1.2 | 0.7 | 2.0 |
|  | 3 | 30 | 155,675 | 14.7% | 18.7 | 11.9 | 25.4 | 0.8 | 0.4 | 1.5 |
|  | 4 (low poverty) | 17 | 69,182 | 6.5% | 23.4 | 12.2 | 34.6 | 1.0 | – | – |
| Hispanic/Latinx | Total | 411 | 1,229,614 | 100.0% | 45.2 | 40.7 | 49.6 |  |  |  |
|  | 1 (high poverty) | 159 | 321,918 | 26.2% | 67.9 | 57.0 | 78.7 | 3.1 | 2.1 | 4.7 |
|  | 2 | 147 | 443,554 | 36.1% | 44.4 | 37.1 | 51.7 | 2.1 | 1.4 | 3.1 |
|  | 3 | 76 | 291,469 | 23.7% | 37.3 | 28.6 | 45.9 | 1.7 | 1.1 | 2.7 |
|  | 4 (low poverty) | 29 | 172,673 | 14.0% | 21.6 | 13.5 | 29.7 | 1.0 | – | – |

Lower bounds for census tract poverty rates by quartile: (1) 21.4%; (2) 12.3%; (3) 6.6%; (4) 0.0%.

**Table 1c.** Mortality due to COVID-19 in Cook County, Illinois by race/ethnicity and census tract poverty rate (Non-Hispanic Black, Non-Hispanic white, and Hispanic/Latinx persons). Ages ≥65, population in households.

| Population group | Census tract poverty quartile | COVID-19 deaths (n) | Population size (N) | % of population group residing in quartile | Adjusted mortality rate |  |  | Mortality rate ratio |  |  |
| --- | --- | --- | --- | --- | --- | --- | --- | --- | --- | --- |
|  |  |  |  |  | Rate per 100,000 | 95% CI (lower bound) | 95% CI (upper bound) | Rate ratio (vs. quartile 4) | 95% CI (lower bound) | 95% CI (upper bound) |
| All three racial/ethnic groups | Total | 1453 | 670,964 | 100.0% | 216.5 | 205.4 | 227.6 |  |  |  |
|  | 1 (high poverty) | 524 | 138,616 | 20.7% | 390.0 | 356.5 | 423.4 | 3.7 | 3.2 | 4.3 |
|  | 2 | 429 | 152,967 | 22.8% | 284.2 | 257.4 | 311.1 | 2.7 | 2.3 | 3.2 |
|  | 3 | 281 | 177,666 | 26.5% | 157.5 | 139.1 | 175.9 | 1.5 | 1.3 | 1.8 |
|  | 4 (low poverty) | 219 | 201,715 | 30.1% | 105.3 | 91.3 | 119.2 | 1.0 | – | – |
| White, non-Hispanic | Total | 490 | 418,071 | 100.0% | 112.5 | 102.6 | 122.5 |  |  |  |
|  | 1 (high poverty) | 52 | 27,854 | 6.7% | 183.4 | 133.4 | 233.4 | 2.2 | 1.6 | 3.0 |
|  | 2 | 137 | 77,630 | 18.6% | 171.5 | 142.7 | 200.3 | 2.1 | 1.6 | 2.6 |
|  | 3 | 143 | 132,220 | 31.6% | 104.5 | 87.3 | 121.6 | 1.3 | 1.0 | 1.6 |
|  | 4 (low poverty) | 158 | 180,367 | 26.9% | 83.1 | 70.1 | 96.1 | 1.0 | – | – |
| Black, non-Hispanic | Total | 591 | 168,275 | 100.0% | 371.1 | 341.0 | 401.2 |  |  |  |
|  | 1 (high poverty) | 349 | 88,015 | 52.3% | 419.4 | 375.1 | 463.7 | 1.7 | 1.1 | 2.5 |
|  | 2 | 141 | 44,546 | 26.5% | 337.6 | 281.3 | 393.9 | 1.4 | 0.9 | 2.1 |
|  | 3 | 75 | 25,389 | 15.1% | 311.7 | 240.8 | 382.6 | 1.3 | 0.8 | 2.0 |
|  | 4 (low poverty) | 26 | 10,325 | 6.1% | 248.3 | 151.0 | 345.7 | 1.0 | – | – |
| Hispanic/Latinx | Total | 372 | 84,618 | 100.0% | 451.6 | 404.7 | 498.6 |  |  |  |
|  | 1 (high poverty) | 123 | 22,747 | 26.9% | 545.2 | 444.4 | 646.1 | 1.6 | 1.1 | 2.4 |
|  | 2 | 151 | 30,791 | 36.4% | 506.0 | 423.5 | 588.5 | 1.5 | 1.0 | 2.2 |
|  | 3 | 63 | 20,057 | 23.7% | 326.7 | 245.1 | 408.3 | 1.0 | 0.6 | 1.5 |
|  | 4 (low poverty) | 35 | 11,023 | 13.0% | 333.2 | 220.3 | 446.1 | 1.0 | – | – |

Lower bounds for census tract poverty rates by quartile: (1) 21.4%; (2) 12.3%; (3) 6.6%; (4) 0.0%.

The adjusted COVID-19 mortality rate for the total study population (N = 4,762,460) was 49.0 per 100,000 (95% CI: 47.1, 51.0). While the NHB population experienced the greatest number of deaths and highest crude mortality rate, its adjusted mortality rate (77.1 per 100,000; 95% CI: 72.0, 82.2; Figure 1) was lower than that of the Hispanic/Latinx population (135.1 per 100,000; 95% CI: 118.1, 152.1). The NHW population had the lowest adjusted mortality rate, 22.8 per 100,000 (95% CI: 21.0, 24.5). Rate ratios comparing adjusted mortality rates to NHWs were 3.4 (95% CI: 3.1, 3.7) for NHBs and 4.5 (95% CI: 4.0, 5.0) among Hispanic/Latinx people.

**Figure 1.**
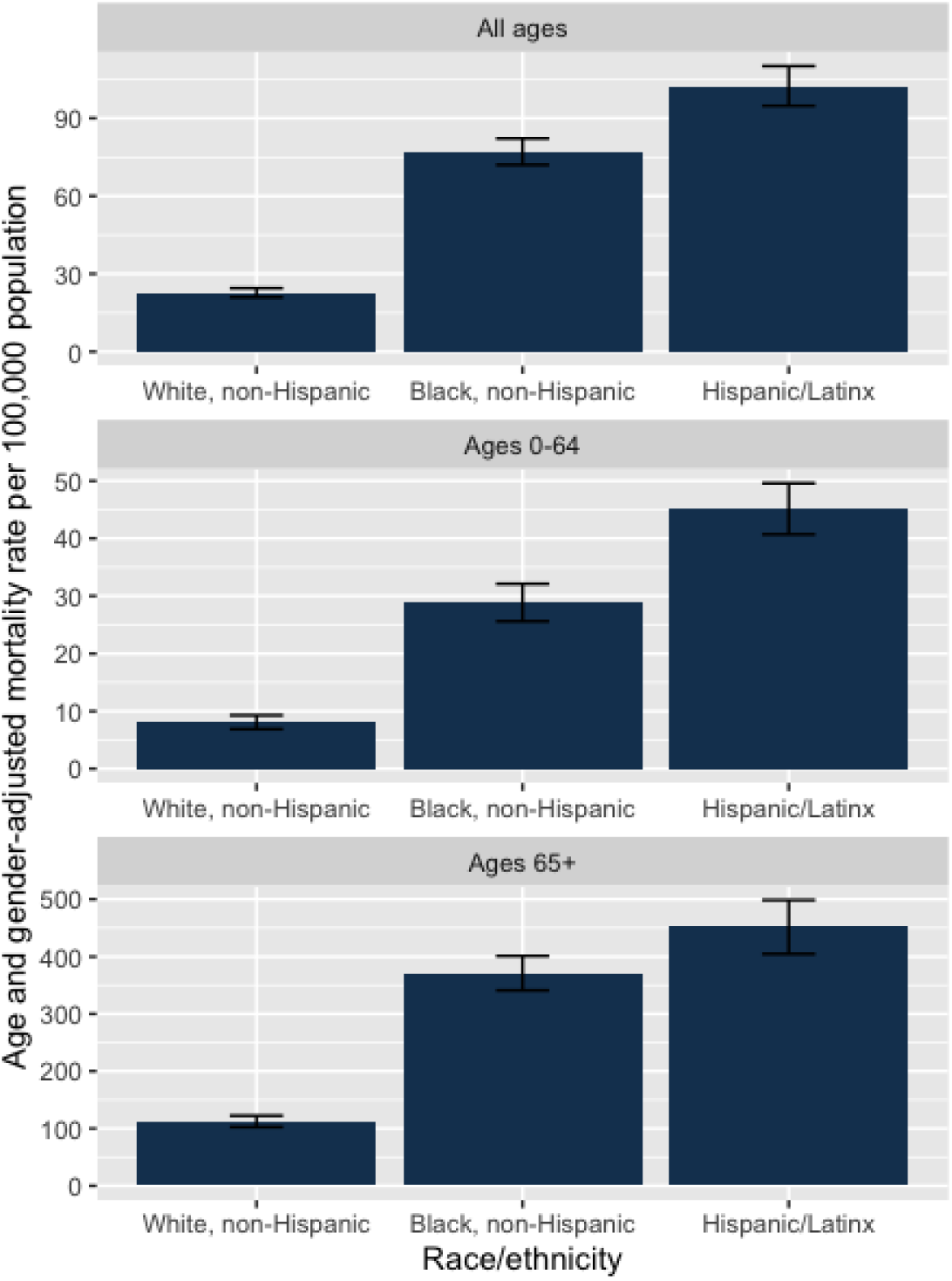
COVID-19 mortality rates by race/ethnicity and age group for the population in households (Cook County, Illinois)

A majority of the NHB population, 54.5% (n = 670,615), resided in the highest-poverty census tract quartile, while 7.7% (n = 171,567) of NHWs lived in these high-poverty areas, as did 26.3% (n = 344,665) of the Hispanic/Latinx population (Figure 2). A plurality of NHWs (42.0%; n = 930,829) resided in the lowest-poverty quartile, as did 6.5% (n = 79,507) of the NHB population and 14.0% (n = 183,696) of the Hispanic/Latinx population.

**Figure 2.**
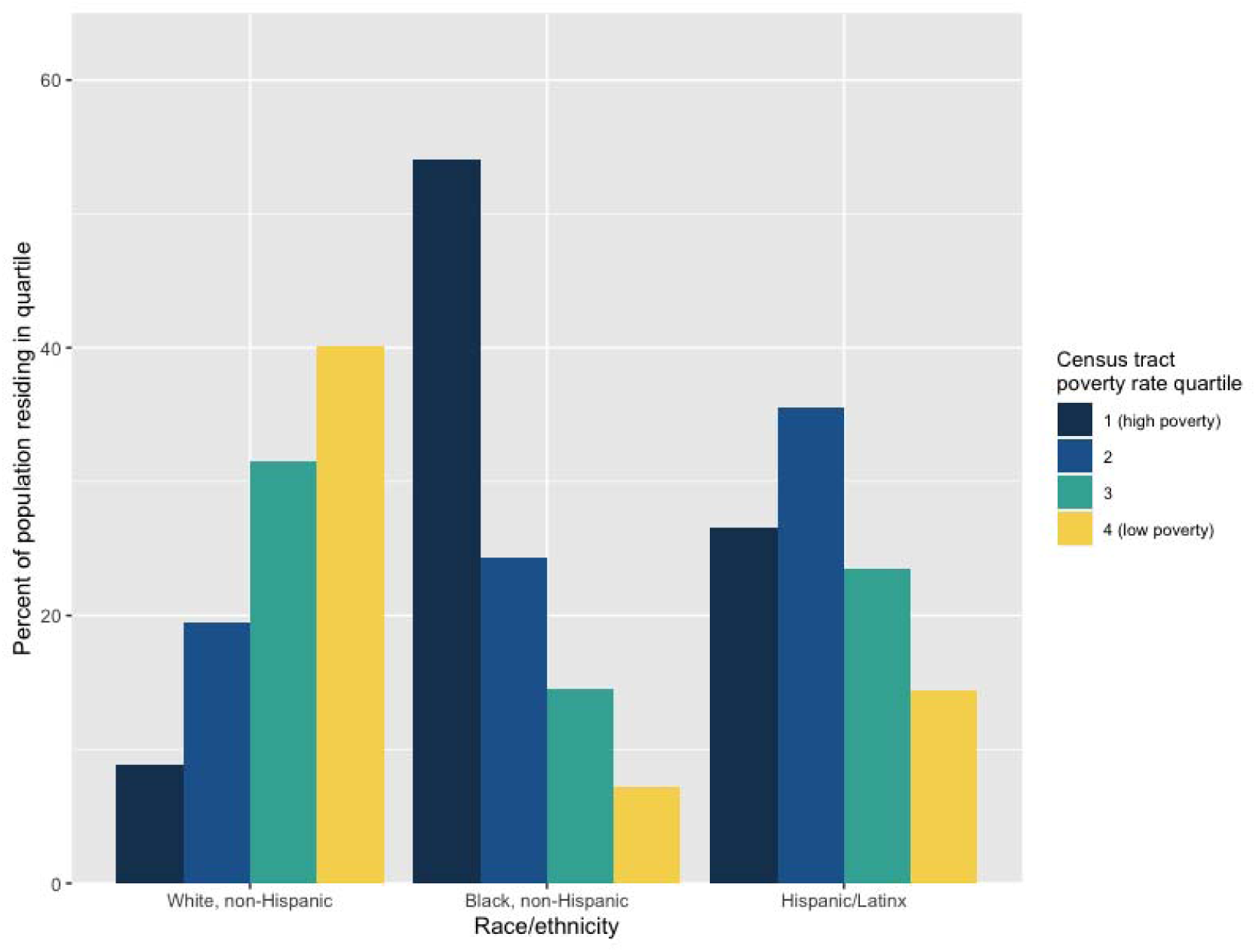
Population distribution of census tract poverty quartiles (Cook County, Illinois, 2018 American Community Survey Data)

Rate ratios comparing mortality in the highest-versus lowest-poverty quartiles within racial/ethnic groups of all ages were: 4.1 (95% CI: 3.3, 5.2) among NHWs, 1.6 (95% CI: 1.2, 2.2) for the NHB population, and 2.1 (95% CI: 1.5, 2.8) for the Hispanic/Latinx population. Socioeconomic gradients were present within all three racial/ethnic groups at both age ranges (Figure 3). For the younger age group (<65 years), the mortality rate for NHWs in the highest-poverty quartile was 13.5 times that of NHWs in the lowest-poverty quartile (95% CI: 8.5, 21.4). Younger NHWs in the highest-poverty quartile had an adjusted mortality rate (37.8 per 100,000; 95% CI: 28.0, 47.6) similar to that of younger NHBs in the highest-poverty quartile (33.1 per 100,000; 95% CI: 28.2, 38.1). In contrast, younger NHBs and Hispanic/Latinx people in the lowest-poverty quartile had mortality rates nearly three times that of younger NHWs in the same quartile. For the older population (≥65 years) the mortality rate among NHWs in the highest-poverty quartile was less than that of even the lowest-poverty NHB and Hispanic/Latinx subgroups.

**Figure 3.**
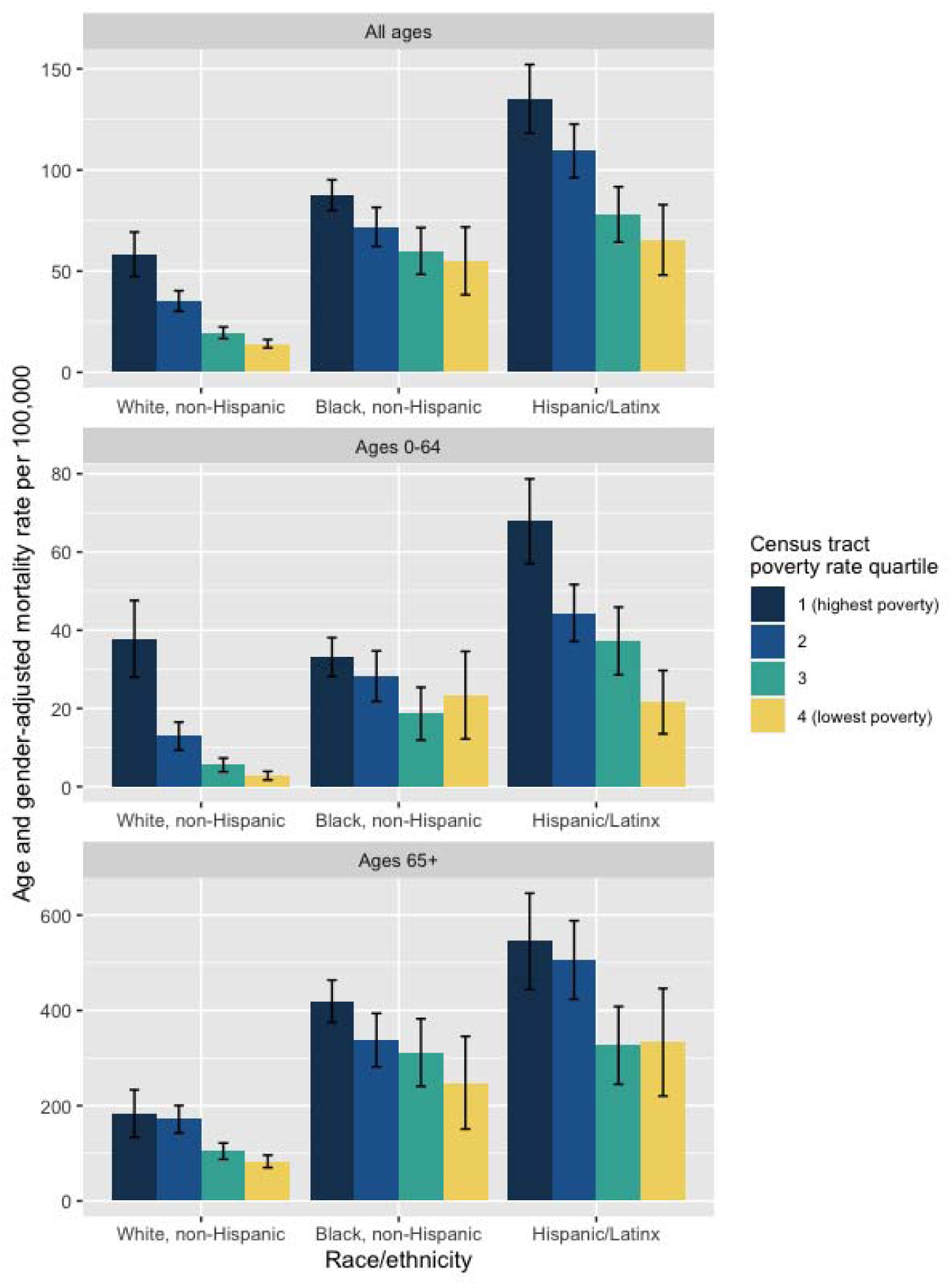
COVID-19 mortality by race/ethnicity, census tract poverty, and age group for the population in households (Cook County, Illinois)

## Discussion

Our analysis of household-related COVID-19 mortality in Cook County, Illinois identified new evidence of economic inequities within racial/ethnic groups. At both younger (<65 years) and older (≥65 years) ages, COVID-19 mortality rates among NHWs, NHBs, and Hispanic/Latinx people exhibited social gradients by census tract poverty quartile. In the highest-poverty neighborhoods, younger NHWs and NHBs had similar mortality rates. In the lowest-poverty neighborhoods, however, younger NHBs and Hispanic/Latinx people had far higher rates than younger NHWs. For the older population, NHWs in the highest-poverty quartile were less likely to die of COVID-19 than even NHB and Hispanic/Latinx people in the lowest-poverty quartile.

One limitation of our study is that we were not able to remove the population residing in institutions from the denominators in rate calculations because census tract-level data were unavailable in the American Community Survey. However, census data for the state of Illinois show that <7% of people ages ≥75 years reside in institutions, suggesting that any resulting bias is likely to be small. Additionally, while the use of area-based poverty measures has been previously validated as a proxy for individual-level socioeconomic position, persons of high socioeconomic position may live in high-poverty neighborhoods and vice versa.

Our findings suggest racial/ethnic inequalities in COVID-19 mortality are partly, but not entirely, attributable to the higher average socioeconomic position of NHWs relative to the NHB and Hispanic/Latinx populations. Within poverty quartiles, there may be persistent racial/ethnic differences in COVID-19 risk factors for exposure (e.g. due to occupational hazards and residential crowding) and infection fatality rates (e.g. due to comorbidities and health care access/quality). Future research on health equity in COVID-19 outcomes should collect and analyze individual-level data on the potential mechanisms driving population distributions of exposure, severe illness, and death. In many US jurisdictions, death certificates already contain useful data on educational attainment and employment by occupation and industry, but these have not yet been made publicly available for the time period of the COVID-19 pandemic. Such data, available in a timely manner, would permit better understanding of these observed patterns and help guide prevention strategies.

## Data Availability

Data available on request to the author.

